# Curbing the AI-induced enthusiasm in diagnosing COVID-19 on chest X-Rays: the present and the near-future

**DOI:** 10.1101/2020.04.28.20082776

**Authors:** Alexandru Burlacu, Radu Crisan-Dabija, Iolanda Valentina Popa, Bogdan Artene, Vasile Birzu, Mihai Pricop, Cristina Plesoianu, Daniele Generali

## Abstract

In the current context of COVID-19 pandemic, a rapid and accessible screening tool based on image processing of chest X-rays (CXRs) using machine learning (ML) approaches would be much needed. Initially, we intended to create and validate an ML software solution able to discriminate on the basis of the CXR between SARS-CoV-2-induced bronchopneumonia and other bronchopneumonia etiologies.

A systematic search of PubMed, Scopus and arXiv databases using the following search terms [“artificial intelligence” OR “deep learning” OR “neural networks”], AND [“COVID-19” OR “SARS-CoV-2”] AND [“chest X-ray” OR “CXR” OR “X-ray”] found 14 recent studies. Most of them declared to be able to confidently identify COVID-19 based on CXRs using deep neural networks. Firstly, weaknesses of artificial intelligence (AI) solutions were analyzed, tackling the issues with datasets (from both medical and technical points of view) and the vulnerability of used algorithms. Then, arguments were provided for why our study design is stronger and more realistic than the previously quoted papers, balancing the possible false expectations with facts.

The authors consider that the potential of AI use in COVID-19 diagnosis on CXR is real. However, scientific community should be careful in interpreting statements, results and conclusions regarding AI use in imaging. It is therefore necessary to adopt standards for research and publication of data, because it seems that in the recent months scientific reality suffered manipulations and distortions. Also, a call for responsible approaches to the imaging methods in COVID-19 is raised. It seems mandatory to follow some rigorous approaches in order to provide with adequate results in daily routine. In addition, the authors intended to raise public awareness about the quality of AI protocols and algorithms and to encourage public sharing of as many CXR images with common quality standards.

## I. BACKGROUND

Due to its accessibility and availability, chest X-ray (CXR) imaging is well established and widely used as a screening and diagnostic tool. In the current context of COVID-19 pandemic, there is a growing demand for imaging and diagnostic services. As the workload is rapidly increasing, it can get overwhelming for radiologists to meet the reporting requirements. As the manual analysis of CXR can be time-consuming, (combined with a growing COVID-19 incidence), a machine learning (ML) approach for the screening process would be much needed.

ML has seen some major breakthroughs over the last few years, drawing a lot of attention from the healthcare system. The recent interest of medical image processing domain in exploring deep learning (DL) has led to development of numerous Artificial Intelligence (AI) diagnostic solutions, part of them (at least 15 AI platforms involved in medicine, including 11 in the imaging field) already being approved by the US Food and Drug Administration board in 2018 (1).

Recent studies suggest neural networks (NN) have surpassed even human performance in some visual and auditory recognition tasks, which may portend its applications in medicine and healthcare, especially in medical imaging (2). Despite this huge enthusiasm, there are some uncertainties that should curb this upsurge.

The “*black-box*” issue, firstly spotlighted by Dean Pomerleau in 1991, has become exponentially harder and more urgent (3). It draws attention to the ML methods to find those worthwhile signals in the vast amount of data available, but also to the validity of the results and the trust in DL (3). Despite remarkable performance of AI, the “*black-box*” problem might be worse than anticipated, as DL models can be surprisingly easy to mislead, and also tend to make overconfident predictions (3, 4).

## II. AIMS AND METHODS

Initially, we aimed to create and validate a software solution able to discriminate on the basis of the CXR between bronchopneumonia lesions induced by SARS-CoV-2 and other different bronchopneumonia lesions (e.g. influenza), using AI/ML models for scanning chest radiographs in the emergency department.

At the end of March 2020, we published and registered our protocol with ClinicalTrials.gov Identifier: NCT04313946. We envisioned a multi-centric study (“*Artificial Intelligence Algorithms for Discriminating Between COVID-19 and Influenza Pneumonitis Using Chest X-Rays - AI-COVID-Xr*”) enrolling patients from Italy, United Kingdom and Romania.

For this purpose, the electronic databases of PubMed, Scopus and arXiv were systematically searched for relevant articles from the inception until late March 2020. The search terms used were [“artificial intelligence” OR “deep learning” OR “neural networks”], AND [“COVID-19” OR “SARS-CoV-2”] AND [“chest X-ray” OR “CXR” OR “X-ray”]. After removing the duplicates, screening titles and abstracts, and assessing eligibility of the selected full texts, 14 valid articles were found. Journal articles published with full text or abstracts in English were eligible for inclusion. However, only in the first two weeks of April at least 12 studies were found declaring that deep neural networks techniques can confidently identify COVID-19 based on CXRs (see Table 1, and references (5, 6)).

**Table 1.**
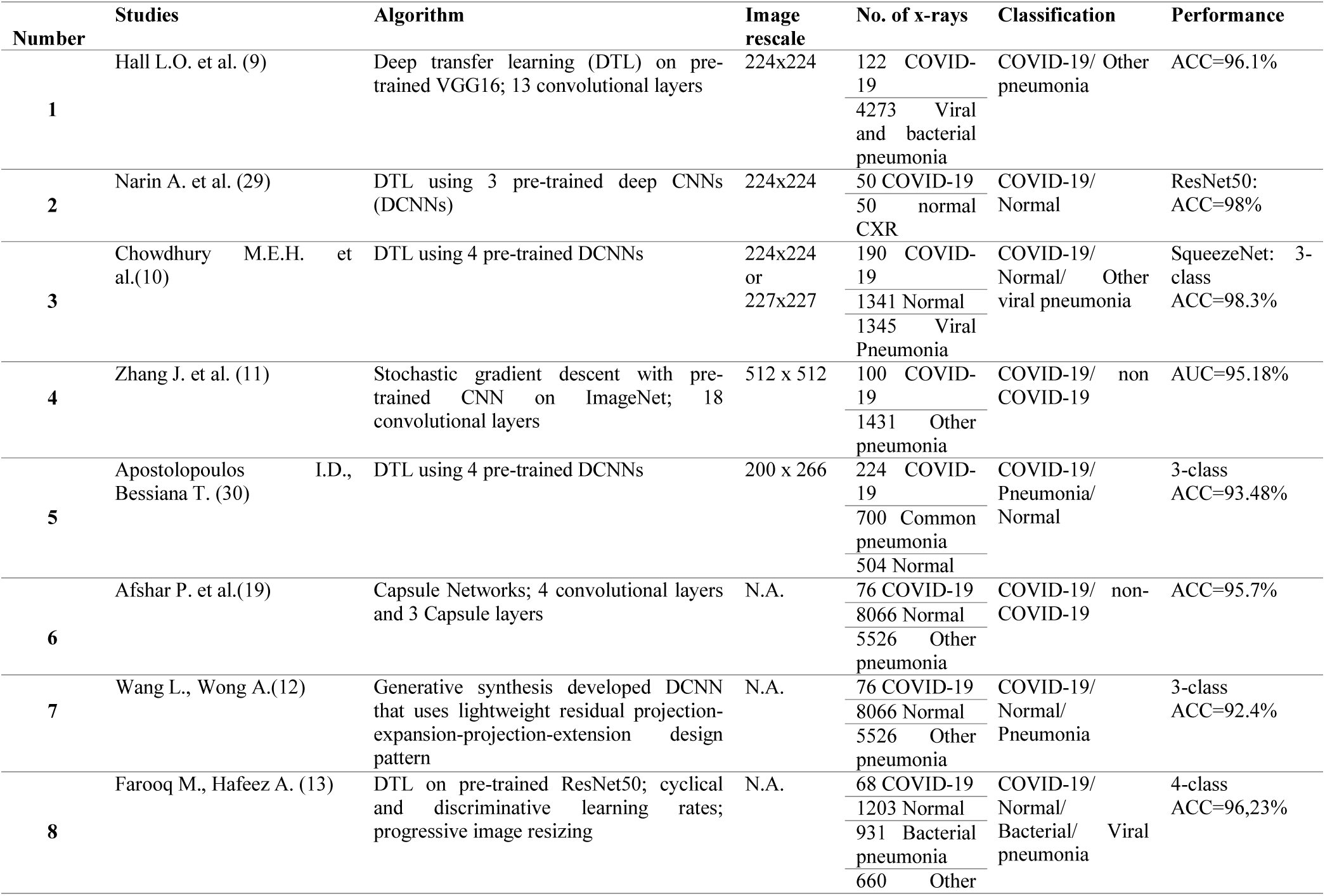
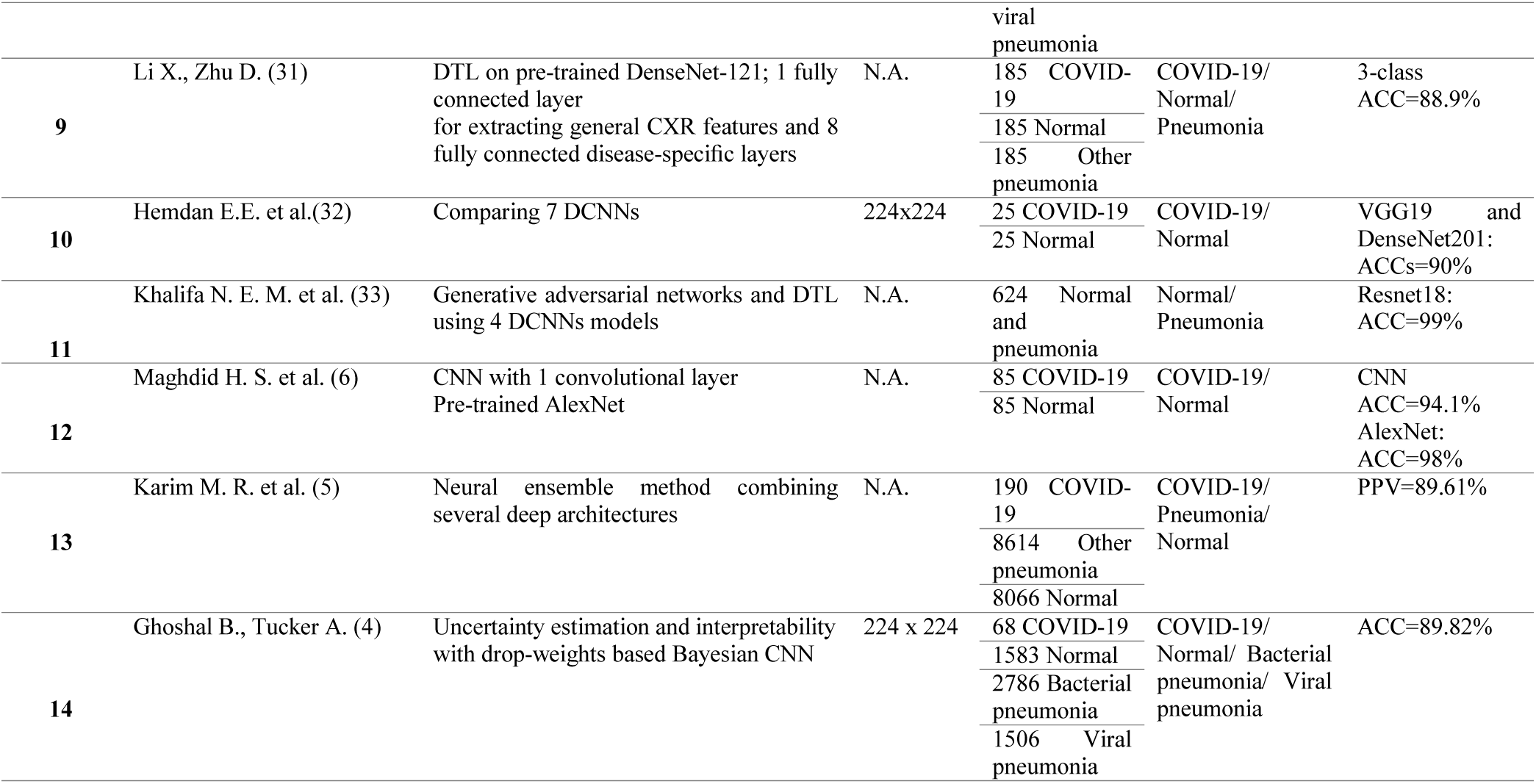
Reported papers using Artificial Intelligence in COVID-19 chest x-ray diagnosis.

Therefore, this paper aims to analyze closer the weaknesses of these AI solutions, trying to cast a glance inside the “*black box*”. Moreover, arguments are also provided supporting our study is realistic with evidence-based findings, avoiding any possible false expectations.

## III. RESULTS AND DISCUSIONS

### RECENT ATTEMPTS, ALLEGED SOLUTIONS: MUCH ADO FOR … (ALMOST) NOTHING?

Most approaches for COVID-19 pneumonia detection on CXRs are based on DL and transfer learning (TL) models. DL and TL represent state-of-the-art performance in image processing and computer vision. Both should integrate the feature detection component and the classification component. TL is a process of adapting a pre-trained model for solving a new problem. The feature detection part is left unchanged and the upper layers corresponding to the classification part are optimized.

#### Testing for biases

Firstly, so far there are no attempts examining whether any of these algorithms, shown in Table 1, claiming to diagnose COVID-19 on chest radiographs *display systematic bias*. In a serious context like the COVID-19 pandemic, when decision-making is vitally important, testing for biases is mandatory. This can be done using different toolboxes (FairML, Lime, Grad-CAM) (7, 8).

#### Issues with the datasets

**a)** All approaches used only public data from covid-chestxray-dataset (https://github.com/ieee8023/covid-chestxray-dataset) and kaggle competitions datasets related to pneumonia. Data owners warn users to *not claim diagnostic performance without a clinical study*.

**b)** CXRs from these public datasets *are not evaluated from a medically point of view*. Each radiograph projection must be correctly identified as PA (postero-anterior) and AP (antero-posterior). Although most films are PA (with the patient in an erect position), in the context of a major outbreak, for severe cases, AP projection will be used for patients confined in bed, in a supine position. This differentiation is important because, for example, in a supine film, blood will flow more to lungs’ apices than when standing. Omitting to differentiate PA from AP projections may result in a misdiagnosis of pulmonary congestion. Moreover, patients lying in bed are more likely to present obvious opacities such as electrocardiogram electrodes, chest drains, pacemakers or foreign bodies, that can alter program’s output.

Another important quality assessment is checking for *signs of patient’s rotation* (ideally should be minimal). Severe rotation can make diagnosis difficult by altering the normal cardiomediastinal contour. Checking for a good X-Rays *penetration* is also a standard part of assessing quality. Penetration is the degree to which X-rays have passed through the body and is affected by both the duration of exposure and the power of the radiation beam. An under-penetrated film looks diffusely light and anatomical structures, like the vertebrae behind the heart, are eclipsed. Over-penetration makes radiographs look diffusely dark and lung markings are poorly delineated.

**c)** Datasets used *are highly imbalanced*. If the training data contains 75% positive and 25% negative cases, the algorithm will immediately notice that predicting “positive” all the time improves its success rate (4, 5, 9–13). The training process is just the optimization of a loss function, without any data interpretation. Whenever the algorithm finds a shortcut to maximize the success rate, it will use it immediately. Adjustments can be made in the training process for unbalanced data, but they are no longer effective for very high ratios like the ones presented in Table 1. To date, only 200 COVID-19 CXR images are publicly available worldwide, while there are thousands of images depicting normal or bacterial pneumonia radiographs. After study completion, our team will contribute with hundreds of COVID-19 CXRs from more medical centres: Suceava and Iasi (Romania), Cremona (Italy), London (UK).

**d)** There is no knowledge about *the X-ray machines* on which chest radiographs were performed. The digital X-ray machine artifacts differ from those that appear on X-ray film. Even though traditional film X-ray machines are more accessible, they have a higher frequency of artifacts resulted from user error, especially in the preparation and development of the images (14).

**e)** There is *no metadata about patients*’ background, travel history, country of origin or other useful knowledge. Data is crucial if it is paired with enough context to create meaning. *Without context data might become noise* (15). From a radiologist point of view, there is a high diagnosis suspicion of COVID-19 if the presence of multiple, bilateral and peripheral patchy ground glass opacities is associated with epidemiological context. Although these typical radiographic characteristics are present only 70% of time (16, 17) the epidemiological context can add more certainty to diagnosis.

**f)** Data is *not equally distributed* on age groups, race, comorbidities and other patient characteristics. This can generate knowledge gaps and confounders in the data that inherently lead to biased predictions.

**g)** Artificial Intelligence is *not clever enough without qualitative data to rely on*. Data quality can induce discriminatory properties in classifiers, while unmeasured predictive features generate discrimination (18). A cliché frequently used in ML is “*Garbage in, garbage out*”, since clean and accurate data goes a long way toward determining the quality of the results.

#### Issues with algorithms

**a)** Most of the studies on DL-based COVID-19 detection have utilized convolutional NN (CNNs) for image classification. CNNs are powerful image processing techniques but have some downsides in the context of CXR-based pneumonia detection. Different models are either encoded for image classification (VGG, DenseNet, GoogleNet, ResNet, MobileNet) or lesions localization, but not both. However, an accurate CXR diagnosis method needs to have embedded both the lesions localization and diagnostic classification components.

Most of the models referred in Table 1 are trained using the whole image, without any interpretation or lesion location information, having only the classification label (e.g. positive/negative). Classification models do not encode the position and orientation of objects ignoring spatial relations between them. Consequently, CNNs of this type are not invariant to rotations and transformations of input data and will need very large datasets to capture all the possible transformations in order to achieve very good performance. However, only small datasets are yet available for COVID-19 classification (19).

In the present context of COVID-19 imagistic detection, integrating models like Mask-RCNN (20) that take into account the localization of lesions might prove more useful and accurate. Also, the RSNA dataset contains annotations, bounding box on each of the pneumonia opacities (https://www.rsna.org/en/education/ai-resources-and-training/ai-image-challenge/RSNA-Pneumonia-Detection-Challenge-2018).

**b)** Most of the models are based only on DL and TL. Even though DL algorithms are very powerful they need to be fed with much larger and qualitative datasets for the network to learn correctly the distinctive diagnostic features. For now, DL may not be the solution and the widespread concept that DL is the only ML method worth investigating could be untrue. Other baseline models with similar performance but with more explainability might prove to be a wiser choice in a context of small-scale datasets available (21).

**c)** Most images in the referred papers were rescaled to a resolution of 224×224 before being used for the DL models training. The information might be present in the shrunken images, but it would be more reliable if the images could be processed at their (higher) initial resolution.

### A NOVEL APPROACH OF AI-ALGORITHMS IN COVID-19 RECOGNITION ON X-RAYS

To our knowledge, no clinical study for COVID-19 CXR diagnosis was conducted before. For more accurate results, we propose rigorous criteria to be followed in the context of a prospective clinical trial.

Firstly, care must be taken for performing good quality radiographs and a more rigorous selection of proper images, equally distributed on age groups. A prospective study allows for better compliance monitoring on the quality of this step.

All images must be subjected to the preprocessing stage. Data normalization to the same range of grays, resolution and zoom is required. All images must have the same mean gray value. Sharpening, histogram equalization (HGE) and Perona-Malik diffusion may be performed for edge enhancement, contrast enhancement and noise reduction on entire CXR images (22). HGE can attain almost equally distributed intensities through blending gray-levels with low frequencies into one and expanding frequent intensities over a higher range of gray levels (5).

Segmentation is another essential step in AI-based COVID-19 image processing and analysis. It outlines the CXR regions of interest (lungs, diseased regions and lesions) in order to feed the ML algorithm with localization information and more meaningful data. Many publications consider segmentation as an essential stage in analyzing COVID-19 images. However, works involving segmentation in COVID-19 are only conducted on CT imaging studies (23).

Segmentation of X-ray images is difficult due to the overlapping of the ribs and clavicles with lesions of interest. There are no methods developed, to date, for segmenting X-ray images of COVID-19 patients. Still, there are several models proposed for CXR segmentation in pneumonia (24–26).

Data enrichment is a pivotal step for bringing context into data. Context is used by the algorithms to discriminate the noise from meaningful data. Table 2 shows main history, clinical and biological variables to be employed in the analysis.

**Table 2.** Data enrichment with context variables from COVID-19 patients.

| **Category** | **Variables** | **Reference** |
| --- | --- | --- |
| **Patient's history** | Age | (34) |
|  | Travel history |  |
|  | Country and city of origin |  |
|  | Comorbidities |  |
| **Clinical data** | Fever, cough, fatigability | Fever and cough are the most frequent symptoms<br>(35, 36) |
|  | Blood pressure, Respiratory rate, dyspnea, oxygen saturation | (37) |
| **Biological data** | Leucocyte, lymphocyte, neutrophils and platelet counts | Lymphopenia is common<br>(35) |
|  | Alanine aminotransferase, aspartate aminotransferase | Liver function tests may be elevated<br>(35, 38) |
|  | D-dimer | Commonly elevated<br>(38) |
|  | CRP, ESR | Elevated and appear to trend upward with disease progression (25, 39) |
|  | LDH, ferritin, troponin, CK, CKMB | (35, 36, 38) |

CRP: C reactive protein; ESR: erythrocyte sedimentation rate; LDH: lactate dehydrogenase; CK: creatine kinase; CKMB: creatine kinase-MB.

For ML models training and testing, we propose a step-up approach from simpler algorithmic methods to more complex, while successively integrating both the localization and the classification modules in a composite framework.

Considering the serious context created by the COVID-19 pandemic and the perilous consequences that can be attained by erroneous models that move from paper to product, the use of debiasing methods in algorithm development should be considered. Debiasing techniques have been proposed for medical imaging ML models like unlearning dataset membership (27). Other methods use adversarial learning to generate a debiased input data (28). Also, a fairness check study was carried out on SOTA models that are trained on large public datasets to predict diagnosis from CXR images (18).

## IV. CONCLUSIONS

The authors consider the potential of AI use in COVID-19 diagnosis on CXR is real. However, scientific community should be careful in interpreting statements, results and conclusions regarding AI use in imaging. It is therefore necessary to adopt standard criteria for research and publication of data, as it seems that in the recent months scientific reality suffered manipulations and distortions. Also, a call for responsible approaches to the imaging methods in COVID-19 is raised. It seems mandatory to follow some rigorous approaches in order to provide with adequate results in daily routine. In addition, the authors intended to raise public awareness about the quality of AI protocols and algorithms and to encourage public sharing of as many CXR images with common quality standards.

## Data Availability

Data used to support the findings of this study are available from the corresponding author upon request.

## Registration

ClinicalTrials.gov Identifier: NCT04313946 (https://clinicaltrials.gov/ct2/show/NCT04313946)

